# A Delphi Study to establish consensus on an Australasian minimum dataset for paediatric stroke rehabilitation

**DOI:** 10.1101/2025.11.12.25340122

**Authors:** Frank Muscara, Adam Scheinberg, Catherine O’Sullivan, Suzi Drevensek, Alex Urquart, Sarah Knight, Sam Armstrong, Katie Banerjee, Theresa Carroll, Jimmy Chong, Monica S. Cooper, Kylie French, Jennifer Harben, Andree Innes, Erin Lunn, Eliza Malony, Divyagowri Manila, Riley Moody, Ray Russo, Amy Shaw, Australia and New Zealand Paediatric Stroke Rehabilitation Research Collaborative, Mark T. Mackay

## Abstract

**Background:** This study aimed to develop a consensus-derived minimum dataset (MDS) for childhood stroke to establish an agreed set of tools to measure key functional outcomes identified as important by childhood stroke survivors and their families.

**Methods:** A steering group comprising clinicians of varying rehabilitation disciplines, as well as families with lived experience, was established to oversee the modified Delphi process. Consultations with parents and young people with lived experience determined key functional outcomes of importance to families throughout rehabilitation. Based on this information, a prospective survey-based Delphi process was conducted, to reach consensus for an Australasian MDS for paediatric stroke rehabilitation. An advisory group comprising specialist rehabilitation clinicians from nine rehabilitation services across Australia and New Zealand responded to three surveys, which aimed to reach consensus on the key functional outcomes that are important to measure, the tools to measure these outcomes, and the timepoints for measurement. After each survey, the steering group collated and analysed the data, and confirmed the content of subsequent surveys.

**Results:** Seventy-three participants from 12 rehabilitation disciplines participated. After Survey 3, consensus was reached on the measures for 14 (64%) of 22 functional outcomes in the pre-school age band, and 20 (50%) of the 40 potential measures in the school-aged band. The Canadian Occupational Performance Measure (COPM) was identified as a particularly valuable tool, achieving broad consensus due to its clinical utility, flexibility, and alignment with outcomes of importance to families and clinicians. Agreement on tools to measure other functional outcomes, particularly body function outcomes, reached lower levels of consensus.

**Conclusions:** The agreement on the best measurement tools to be included in a MDS represents an important advancement in paediatric stroke rehabilitation research. The current findings provide a foundation for transformative improvements in future research, and in the care of children recovering from stroke.

## INTRODUCTION

Paediatric stroke is an infrequent occurrence, occurring in 1-13 (1) out of every 100, 000 children per year (2). In adults there is a strong focus on therapies that minimise extent of brain injury by restoring blood flow (3, 4). Children often do not, however, have access to these therapies due to delayed diagnosis, (5–7), leading to significant morbidity and mortality (8), with more than half of survivors experiencing long term impairments, including cognitive, social, motor and language deficits (9, 10).

Rehabilitation is of critical importance in improving functional outcomes (11). There is a paucity of evidence to guide sub-acute stroke care in children (12, 13), with approaches largely extrapolated from adults. The main issues with extrapolating data from adult rehabilitation studies is that the aetiologies of childhood stroke are much more varied with the other major variable being the developmental context of the stroke occurs (14). This greatly impacts on the ability of clinicians to provide accurate and meaningful prognostic counselling to families, and to provide timely, targeted, evidence-based rehabilitation interventions for children at appropriate ages, in the context of a rapidly developing brain.

There has been a shift towards evidence-based practice in acute paediatric stroke care, with the establishment of guidelines for clinical management (5, 6, 12). These guidelines have been developed, however, in the absence of high-quality studies for rehabilitation practices and interventions (15), meaning the clarity and specificity of available recommendations remains limited. Existing evidence for motor interventions is limited mainly to children or adults with cerebral palsy secondary to perinatal stroke (16–18). In cognitive domains, evidence is limited to memory training strategies to improve cognitive function in highly selected populations (12) or extrapolated from adult stroke and traumatic brain injury populations (19). There are limited reviews and amalgamation of data in the field regarding the long-term outcomes in this population (20), or the best interventions to use to improve these outcomes (19).

There is also limited agreement across the breadth of rehabilitation disciplines regarding the most feasible, reliable and valid tools to assess patient functioning within an inter-disciplinary rehabilitation context. Importantly, there have been some recent initiatives to reach consensus on measures for the evaluation of outcomes in childhood stroke. Feldman and colleagues (21) proposed a comprehensive tool kit of 48 outcome measures for evaluation of outcomes in childhood stroke. This process, which involved an international working group, included a review of measures, but did not engage all end user health professionals providing rehabilitation care. Only three global measures included in the list, two of which are typically administered by neurologists, have been validated in the paediatric stroke population. Furthermore, it is unclear whether the measures assess outcomes that are of importance to children and families affected by stroke.

Due to these knowledge gaps in this rare condition, there is a need to establish an agreed upon minimum data set (MDS), which can be developed to collate higher quality outcome data, and to support the establishment of multi-site collaborations and research trials. As a first step to address this, the Australasian Paediatric Stroke Rehabilitation Research Network was established in 2022, engaging clinicians and researchers across all Australian and one New Zealand paediatric rehabilitation services as well as a national childhood stroke lived experience advisory group (LEAG). The objective of the current study, therefore, was to leverage this network and use a Delphi Consensus process, with the aim of establishing an Australasian paediatric stroke rehabilitation MDS.

This process specifically aimed to create a list of agreed upon tools that can measure functional rehabilitation outcomes of importance to childhood stroke survivors and their families. This MDS will allow all participating sites in this network to collect data in a consistent way, leading to the aggregation of data into an Australian and New Zealand database of paediatric stroke outcomes. This will in turn build a platform to conduct future multi-site rehabilitation interventional trials, which will better inform and support evidence-based approaches in paediatric stroke rehabilitation.

## METHODS

### Study Design

The study was approved by the Royal Children’s Hospital Research Ethics Committee (HREC: 85239).

Figure 1shows the process to conduct the three-stage modified Delphi Consensus process for establishing an Australasian paediatric stroke rehabilitation MDS. The study focussed on outcomes of stroke survivors through childhood and adolescence. Newborns (< 29 days of life) were out of scope because they typically classified as having cerebral palsy.

The Delphi consensus process is an efficient method of capturing a broad set of opinions from peers. It is considered to be a robust tool for determining whether a consensus exists within an expert group, consisting of individuals with knowledge of the subject under study, and for clarifying the agreement which exists within that group (22). It is a well validated mechanism of repeated surveys which removes the inherent group-conformity bias that can occur in face-to-face meetings by maintaining the anonymity of respondents (23). The Delphi method achieves consensus through the use of questionnaires, which are administered over a series of rounds. Results of each round are reviewed and interpreted by a steering group (24), and then fed back to the expert group until consensus is achieved (23).

### Participants and roles

Participants comprised young people and parents with lived experience of childhood stroke, and clinicians and researchers with expertise in childhood stroke rehabilitation (see Appendix 1for participating rehabilitation services).

#### Expert Steering Group

An expert steering group was established to provide governance oversight to the initiative. This group comprised of fourteen rehabilitation clinicians. Each rehabilitation site, and each rehabilitation clinician discipline was represented. Three clinical researchers with expertise in paediatric and adult stroke rehabilitation, and three representatives with lived experience were also part of the group. The group’s role also included the analysis and interpretation of the survey data, and to determine the content of subsequent surveys.

#### Advisory Group

The Advisory group was defined as the respondents to the three Delphi survey rounds. To be eligible for inclusion in the group, participants were required to be current clinicians within participating rehabilitation services. These included clinicians from the range of professions that contribute to the rehabilitation process, across all nine Australian paediatric rehabilitation services, which provide care to children in 6 of 8 states and territories, and one service in Auckland, New Zealand. Disciplines from each participating site included: rehabilitation physicians, neurologists, neurosurgeons, nurses, physiotherapists, occupational therapists, speech pathologists, clinical psychologists, neuropsychologists, social workers, teachers or education experts, dieticians, play therapists, music therapists, and allied health assistants.

#### Lived-Experience Advisory Group (LEAG)

Young people and caregivers of young people with lived experience of arterial ischaemic stroke (AIS) or haemorrhagic stroke (HS) were invited to provide consultation to the study. To be eligible, young people were required to have experienced a previous childhood stroke but be currently 16 years of age or older. They were also required to have been treated by a participating rehabilitation site. Parents were required to have a child who experienced a stroke. Participants provided signed, informed consent, and parents of young people aged under 18 years also provided signed, informed consent for their child to participate.

### Theoretical Framework

The theoretical underpinnings of the current study used to inform data collection were based on the International Classification of Functioning, Disability and Health (ICF) Framework (25). Consistent with the ICF, the functional outcomes discussed in the surveys were divided into six domains: Health Condition, Body Structure and Function, Activity, Participation, Personal Factors, and Environmental Factors.

### Procedure for Data Collection and Analysis

A study champion was nominated by each rehabilitation service, to inform the clinicians within that service about the project. The site champions also distributed study information and recruitment flyers through their networks in order to recruit parents and young people with lived experience. The research team contacted potential participants (clinicians, parents and young people) via email and invited them to participate in the study.

After the participants were recruited, LEAG members were invited to provide consultation to the project in groups or individually. Discussions within this consultation focussed on key functional outcomes that LEAG members believed were important for clinicians to measure throughout the rehabilitation journey. The themes emerging from the consultations were used by the Expert Steering Group to inform the content of the Delphi surveys. The Advisory group was then invited to complete three semi-structured surveys which included a number of free-text responses, to gather information about functional outcomes of importance to measure during rehabilitation, and the most appropriate tools and timepoints to measure these outcomes. Results from each survey were collated and analysed by the Expert Steering Group, with these results used to inform the content of each subsequent survey, in order to move toward consensus (process shown in Figure 1).

**Figure 1:**
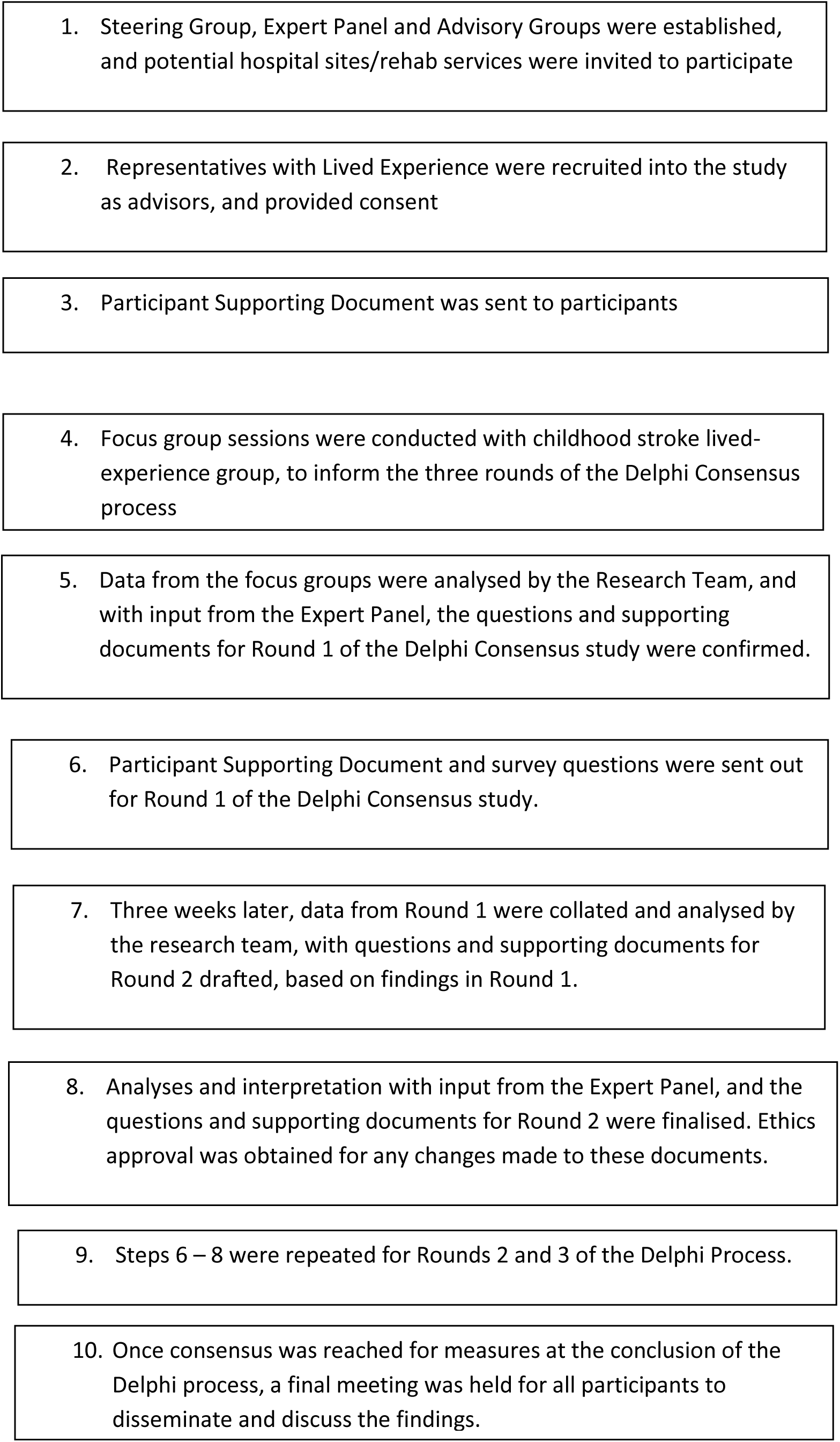
Design and Flow of the Delphi Consensus Process.

#### Process for LEAG Consultation

Health consumers were asked the following questions:

1. What are the most important things for clinicians to measure and monitor in the short term and long-term following a child’s stroke?
2. What are the most appropriate timepoints for clinicians to assess and collect information about these outcomes?
3. How willing would parents be to complete these measurements, and what would be a reasonable length of time for them to spend completing assessment?

A scribe took notes and summarised the key priority areas identified within the interviews and groups, which informed the development of Survey 1 of the Delphi process.

#### The Delphi Consensus Process

Three surveys were sent out to Advisory Group respondents. The Expert Steering Group met after each survey to review and analyse the survey findings. The results of each survey informed the content for subsequent surveys. On completion of three surveys, a final meeting was conducted to inform all study participants of the findings. All meetings were conducted online.

A Decision-Making Guide from the Stroke Recovery and Rehabilitation Roundtable Trials Development Framework (26) was provided to participants to assist with decision making throughout the consensus process, without biasing or influencing their selections (Appendix 2). This framework included the following priority knowledge units:

i. WHAT measures to use for each outcome, considering factors including cost, training and equipment required, validity and reliability, time taken to administer, general feasibility of use for everyone in the network, ability to use remotely and the ability to demonstrate change over time;
ii. WHEN to administer these measures (including the initial and follow up time points;
iii. WHERE? (in the inpatient or outpatient rehabilitation setting); and
iv. WHO was being assessed or treated (including stroke type, and age at assessment). The purpose of this framework was to support clinicians to select the best, most appropriate and feasible measures, rather than their preferred or most commonly used measure.

Surveys were administered and designed using the secure, web-based software platform REDCap (27, 28).

#### Survey 1

Using the ICF framework and the functional outcomes identified in the Australian subacute childhood stroke guidelines (12) as a starting point, the information obtained within the LEAG consultations allowed the generation of a list of functional outcomes that were of importance from the perspective of consumers with lived experience. These functional outcomes were categorised into the domains aligned with the ICF Framework (see Figure 2). Based on feedback from Delphi participants, and following discussion with the Expert Steering Group, motor outcomes were divided into measures that evaluated motor *function*, and measures that evaluated motor *impairment*. Consent forms and the first survey were sent to interested clinicians across the nine participating sites. The survey collected demographic information about the respondents, including their rehabilitation profession and the number of years spent working in that profession, and the three most important factors that they take into consideration when selecting a tool in their clinical practice.

**Figure 2:**
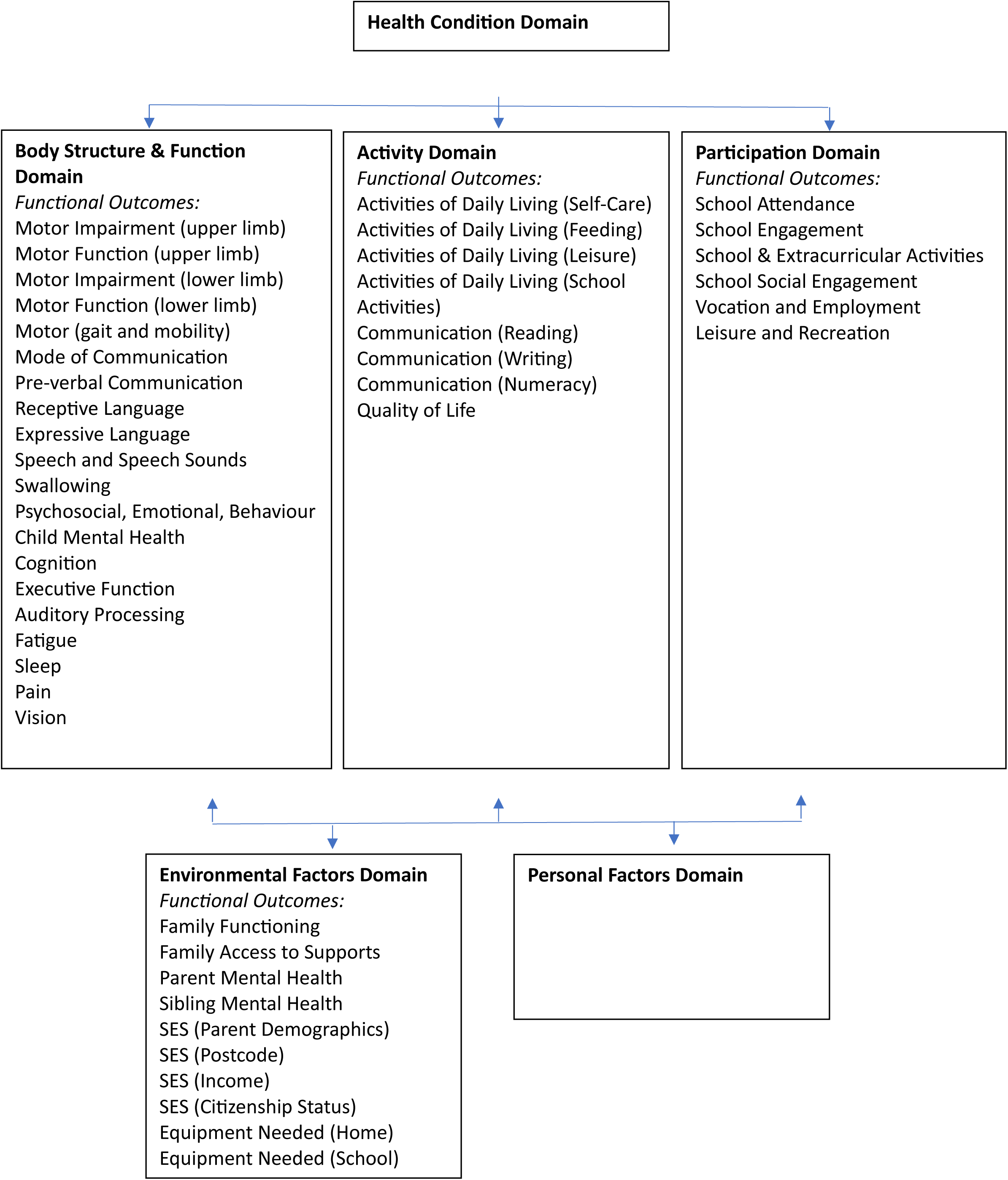
ICF Domains and the proposed Delphi Functional Outcomes within each Domain.

Respondents were invited to critically appraise and rate the importance of the listed functional outcomes on a Likert scale of 1 to 5 (where 1 = Not important at all and 5 = Very important). Clinicians with expertise and experience in specific functional outcomes were asked to list all the tools that would be relevant and appropriate to use to measure the outcome. They were also asked whether the functional outcomes listed were best assessed using an informal approach, such as through interviews or observations, or by using a formal, validated measure.

#### Survey 2

Survey 1 findings were collated and analysed by the Expert Steering Group to inform the content of Survey 2. The list of measures provided by respondents in Survey 1 was reviewed by the Expert Steering Group as well as a group of clinical researchers with expertise in the psychometric properties of the proposed assessment tools. These measures were reviewed and evaluated on factors including validity and reliability, general feasibility of use, and the ability to demonstrate change over time. Following this process, the list of measures provided in Survey 1 was reduced to a select list of measures to be incorporated into Survey 2. In addition, any functional outcomes that reached consensus regarding whether informal assessment was the best measurement approach, were removed from subsequent surveys. Consensus was considered to be reached if more than 70% of respondents selected the informal measurement approach.

In Survey 2, respondents were invited to rank the top two measures from the list arising from Survey 1, for each of ICF domain, where 1 = the best, and 2 = the second-best measure across two age bands, Pre-School (0-5 years) and School aged children (6-18 years). Based on Survey 1 responses, it was decided to split the Pre-School age group into two separate age bands for some measures: Pre-School (0-2 years) and Pre-School (3-5 years). Finally, respondents were asked to indicate the optimal assessment time point for each outcome. Time points post-stroke were based on a rehabilitation recovery road map as outlined by Felling and colleagues (29),with those being: the ‘Acute’ period (0 - 7 days), the ‘Sub-acute’ period (7 days to 6 months), and the ‘Chronic’ period (beyond 6 months). Respondents were asked to indicate which time-point(s) following the stroke event would be optimal in terms of clinical utility to measure these outcomes. More than one period could be selected.

#### Survey 3

Survey 2 findings were collated and analysed by the Expert Steering Group and a group of clinical researchers with expertise in clinical measurement, to inform the content of Survey 3. Consensus was pre-defined as >70% of respondents selecting the same measure. All outcomes that had not yet reached consensus were included in Survey 3. Measures that were ranked highly in Survey 2 or were deemed appropriate, valid and reliable by the Expert Steering Group and the experts in clinical measurement were included in Survey 3.

Respondents were asked to only provide responses for functional outcomes they had expertise with assessing in their normal clinical practice. Respondents were asked to rank the *single* best measure for each functional outcome in each age band.

## RESULTS

### Survey 1

A total of 73 participants consented and responded to Survey 1, including clinicians from 12 disciplines with a wide range of experience (see Table 1). The median length of time they had worked in their current roles was 10.6 years, with a range of 1-30 years.

**Table 1.** Participant demographics (N = 73).

| Rehabilitation Profession<sup>a</sup> (n) |  |
| --- | --- |
| Rehabilitation Physician | 14 |
| Physiotherapist | 14 |
| Occupational Therapist | 11 |
| Speech Pathologist | 8 |
| Neuropsychologist | 6 |
| Social Worker | 4 |
| Nurse | 4 |
| Neurologist | 3 |
| Clinical Psychologist | 3 |
| Teacher/Education expert | 3 |
| Allied Health Assistant | 2 |
| Music Therapist | 1 |

NOTE: <sup>a</sup> These are all specialist paediatric clinicians

Table 2 presents the factors considered by rehabilitation clinicians when selecting a tool or measure. Participants were asked to critically appraise and rate the importance of the listed functional outcomes, within the ICF framework (Table 3). Participants were then invited to list the measures they believed would be the most appropriate, feasible and valid tool to assess each outcome. The full list of measures respondents proposed for each functional outcome are provided in Appendix 3. The refined list of measures included in Survey 2 is in Appendix 4. Consensus was reached for seven functional outcomes, where informal measurement or data collection approaches (such as interviews or observations), were believed to be more appropriate than standardised measures. These findings are displayed in Table 4.

**Table 2.**
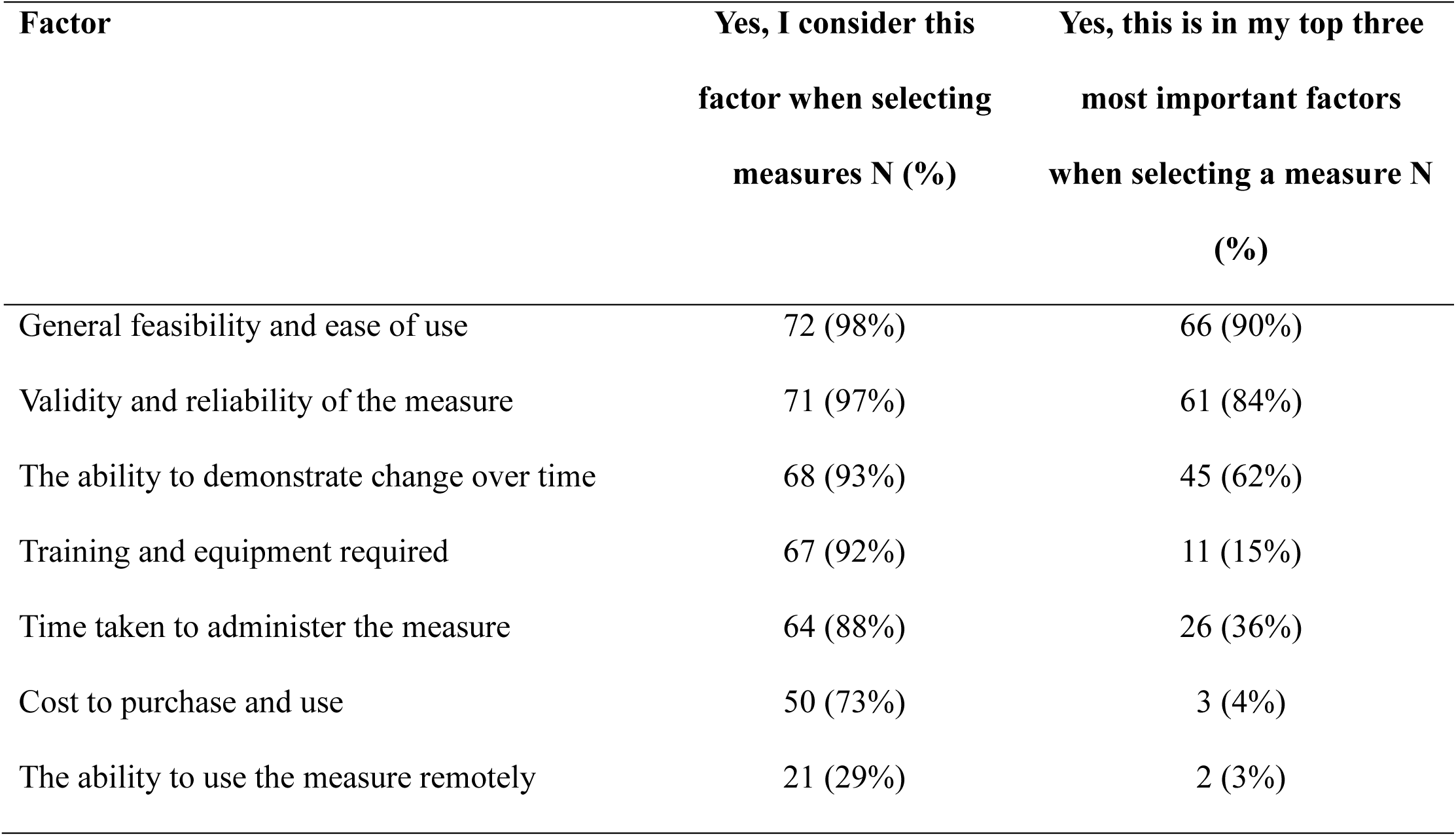
Important Factors Considered by Participants When Selecting Clinical Measures and Tools (N = 73)

| **Factor** | **Yes, I consider this factor when selecting measures N (%)** | **Yes, this is in my top three most important factors when selecting a measure N (%)** |
| --- | --- | --- |
| General feasibility and ease of use | 72 (98%) | 66 (90%) |
| Validity and reliability of the measure | 71 (97%) | 61 (84%) |
| The ability to demonstrate change over time | 68 (93%) | 45 (62%) |
| Training and equipment required | 67 (92%) | 11 (15%) |
| Time taken to administer the measure | 64 (88%) | 26 (36%) |
| Cost to purchase and use | 50 (73%) | 3 (4%) |
| The ability to use the measure remotely | 21 (29%) | 2 (3%) |

**Table 3.** The rated importance of each functional outcome, and whether they should be included in the Minimum Data Set.

| Functional Outcome | Yes, include in the dataset (%) | Importance of measuring this outcome (M)<sup>a</sup> |
| --- | --- | --- |
| *Body Structure and Function Domain* |  |  |
| Motor (upper limb) | 100 | 4.77 |
| Motor (gait and mobility) | 99 | 4.72 |
| Mode of Communication | 99 | 4.76 |
| Motor (lower limb) | 97 | 4.61 |
| Expressive Language | 96 | 4.71 |
| Psychosocial, Emotional, Behaviour | 96 | 4.58 |
| Vision | 94 | 4.70 |
| Receptive Language | 93 | 4.69 |
| Swallowing – Mechanics | 93 | 4.57 |
| Pain | 91 | 4.57 |
| Auditory Processing | 89 | 3.75 |
| Executive Function | 88 | 4.46 |
| Child Mental Health | 87 | 4.51 |
| Pre-Verbal Communication | 86 | 4.46 |
| Cognition | 86 | 4.56 |
| Fatigue | 85 | 4.37 |
| Speech and Speech Sounds | 78 | 4.22 |
| Sleep | 76 | 4.27 |
| *Activity Domain* |  |  |
| Activities of Daily Living (Self-Care) | 97 | 4.85 |
| Activities of Daily Living (Feeding) | 91 | 4.57 |
| Activities of Daily Living (Leisure) | 88 | 4.38 |
| Rehabilitation Goals Attainment | 86 | 4.67 |
| Quality of Life | 85 | 4.54 |
| Activities of Daily Living (School Activities) | 77 | 4.35 |
| Communication – Reading | 73 | 4.22 |
| Communication – Numeracy | 68 | 3.96 |
| Communication – Writing | 63 | 3.94 |
| *Participation Domain* |  |  |
| School - Social | 87 | 4.48 |
| Vocation and Employment | 82 | 4.00 |
| School Attendance | 79 | 4.21 |
| Leisure and Recreation (Family) | 77 | 4.18 |
| School Engagement | 74 | 4.13 |
| School - General Activities | 66 | 3.75 |
| *Environmental Factors Domain* |  |  |
| SES Parent demographics | 95 | 4.49 |
| SES Postcode | 92 | 4.20 |
| SES Citizenship Status | 90 | 4.28 |
| Family Function | 88 | 4.36 |
| Parent Mental Health | 85 | 4.41 |
| Equipment needed (Home) | 82 | 4.41 |
| Equipment needed (School) | 75 | 4.31 |
| Family Access to Supports | 74 | 4.45 |
| SES Parent income | 72 | 3.85 |
| Sibling Mental Health | 59 | 3.87 |

NOTE: <sup>a</sup> Mean rating of a Likert scale from 1 to 5, where 1 = Not important at all and 5 = Very Important); SES = socioeconomic status

**Table 4.**
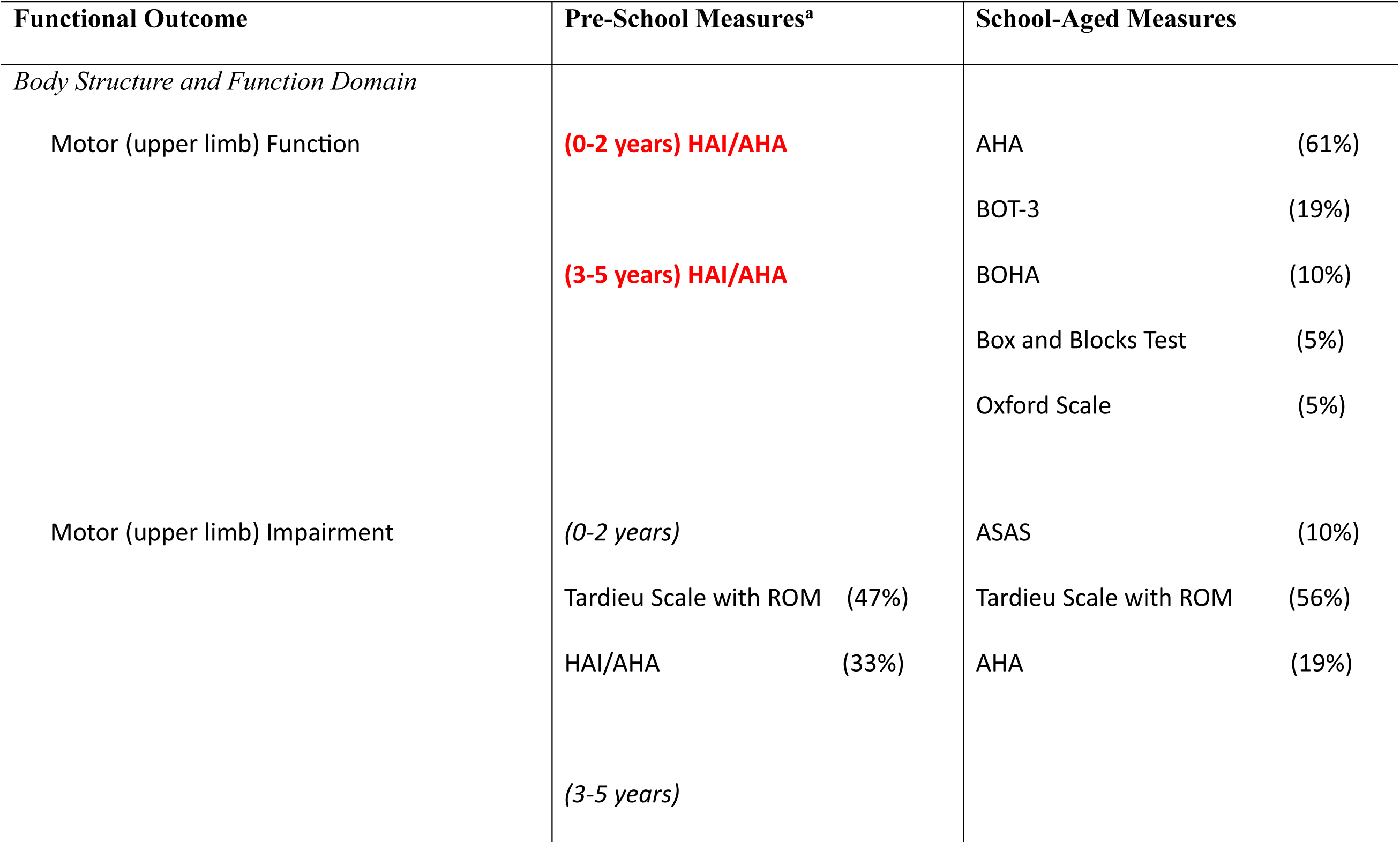
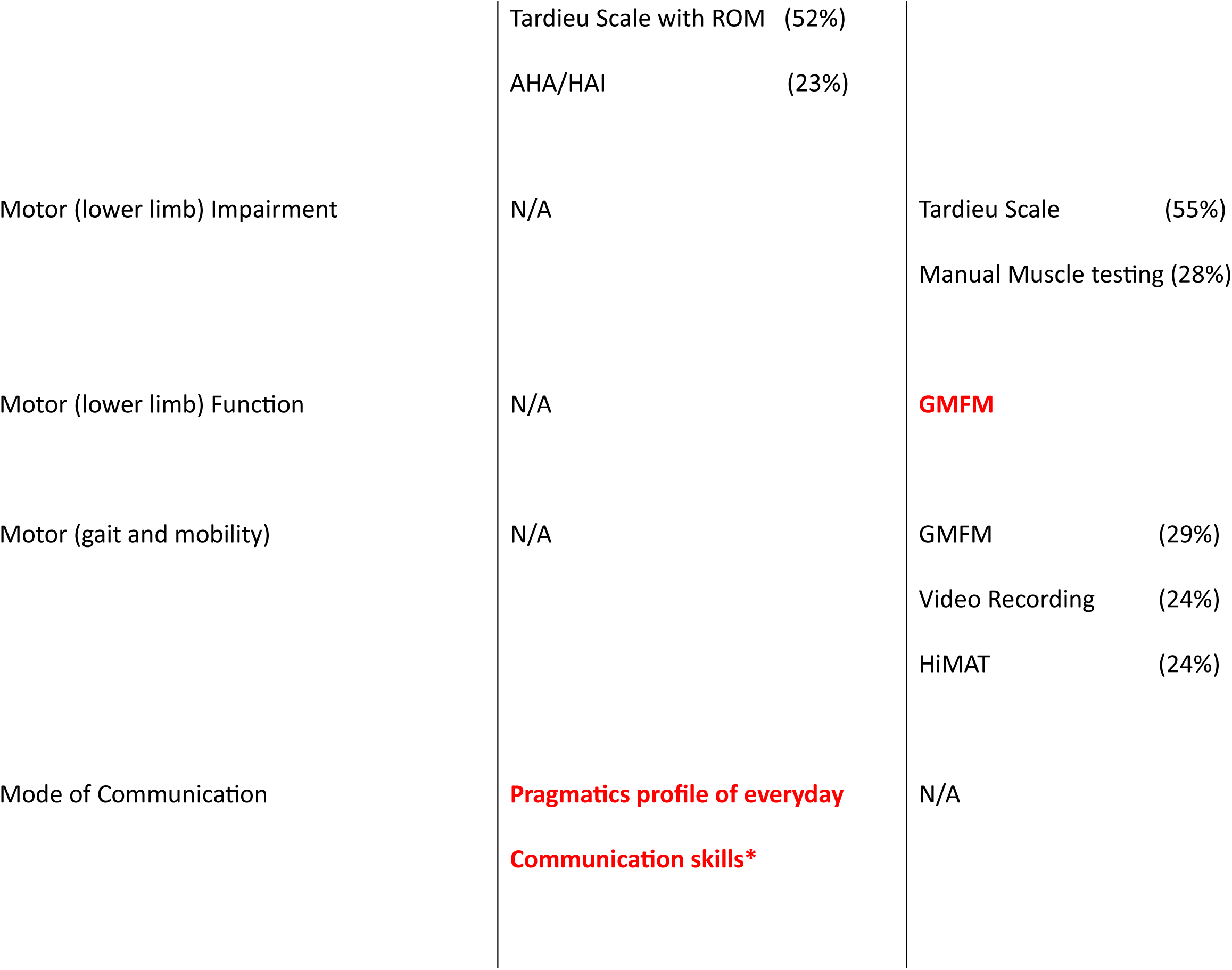
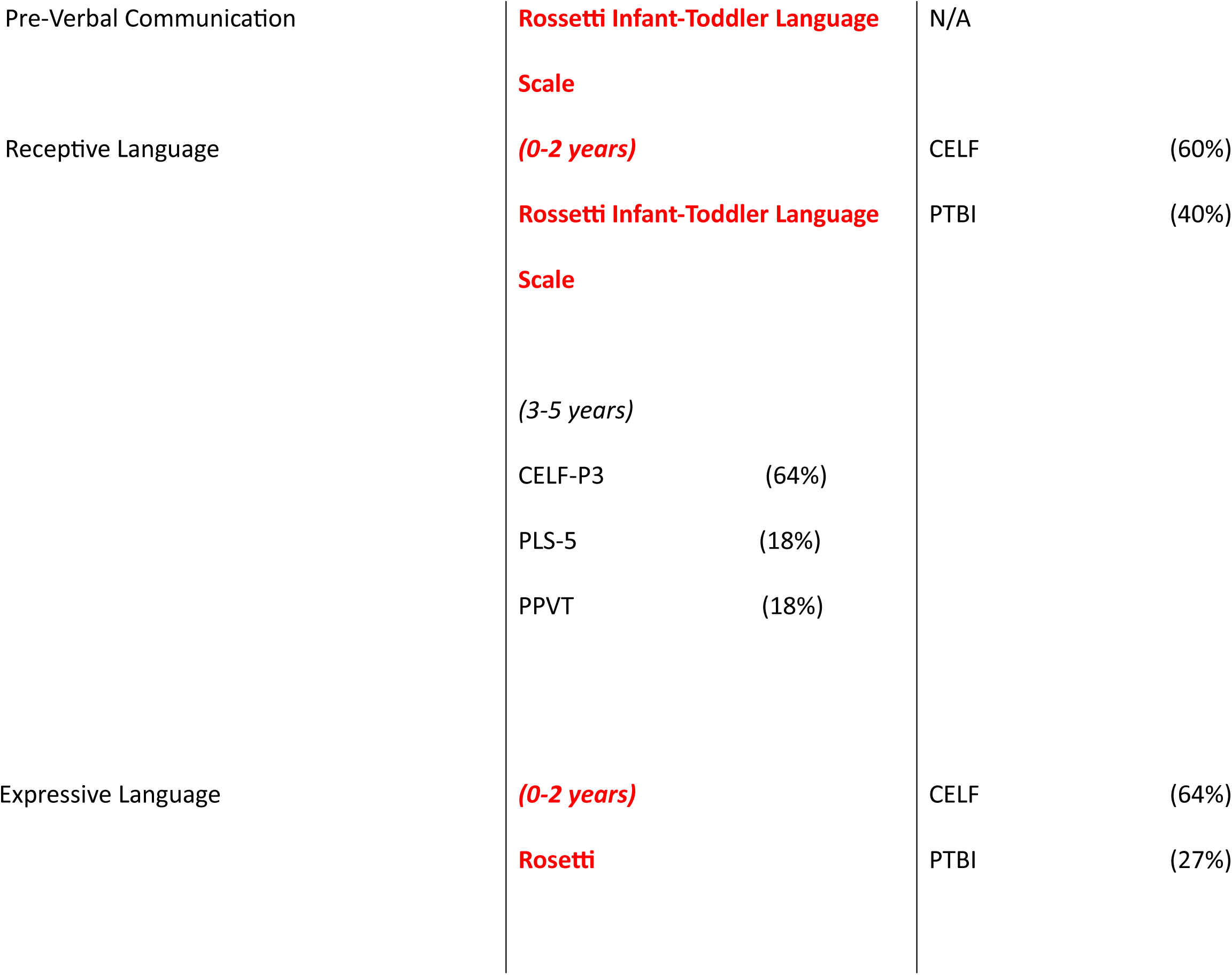
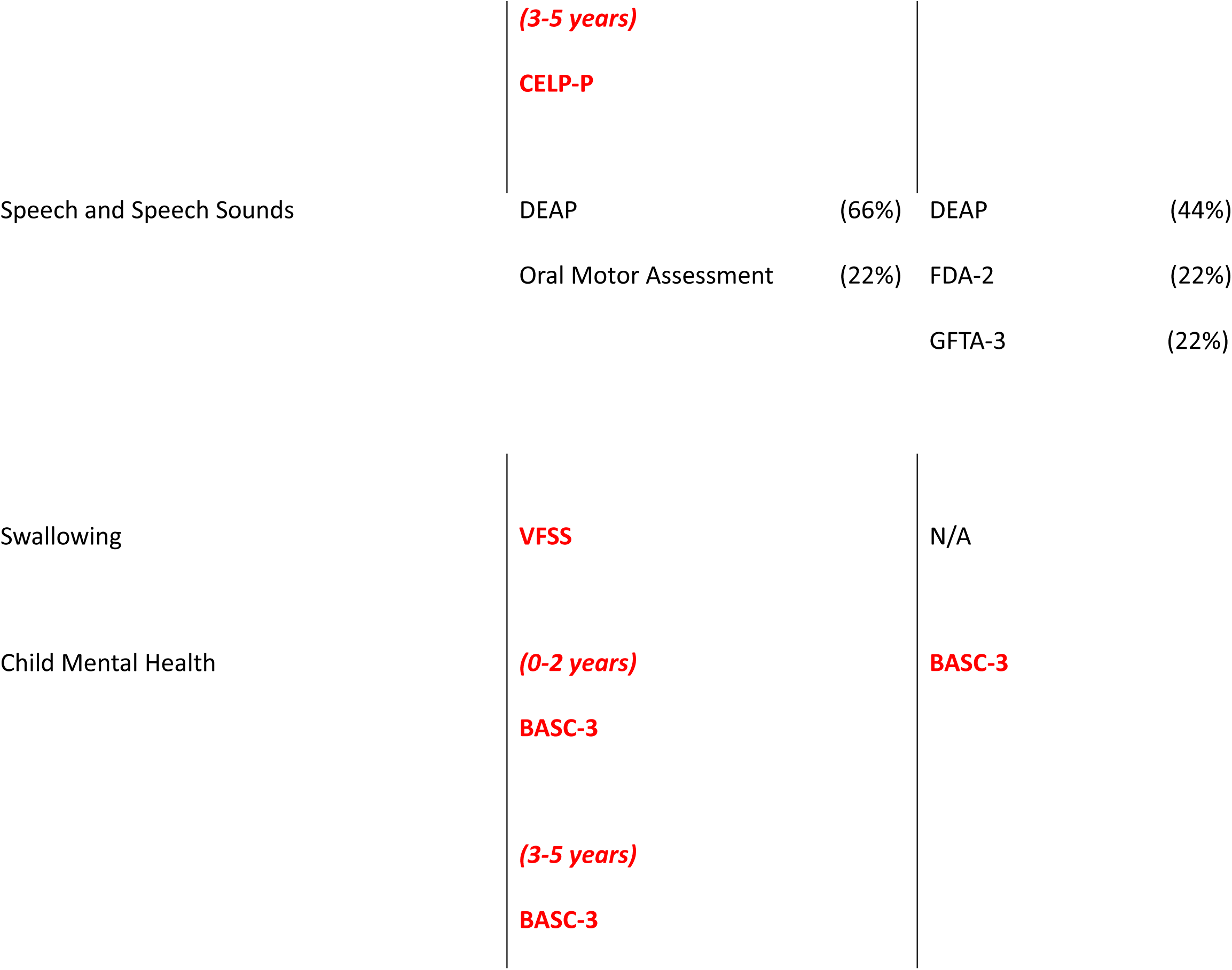
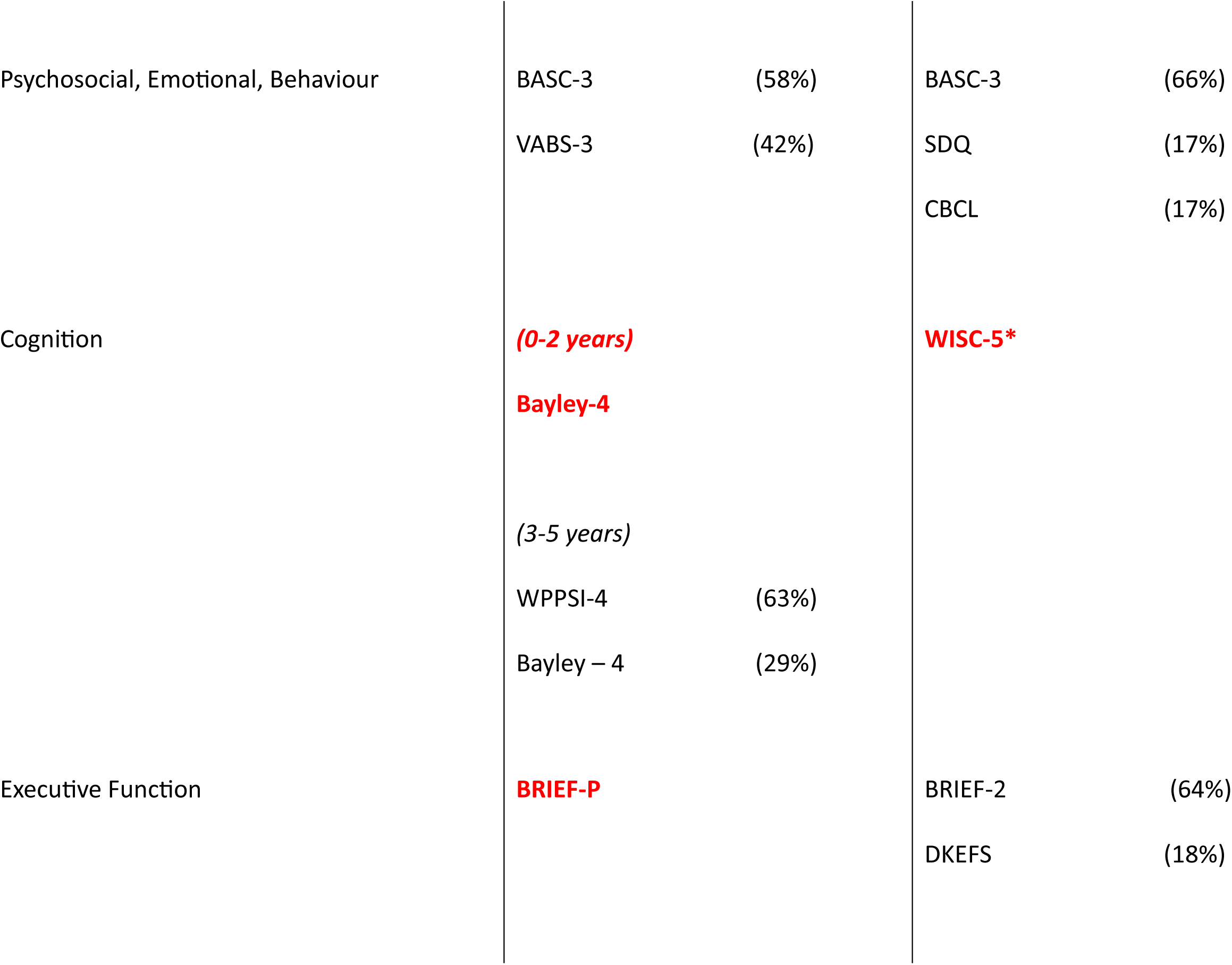
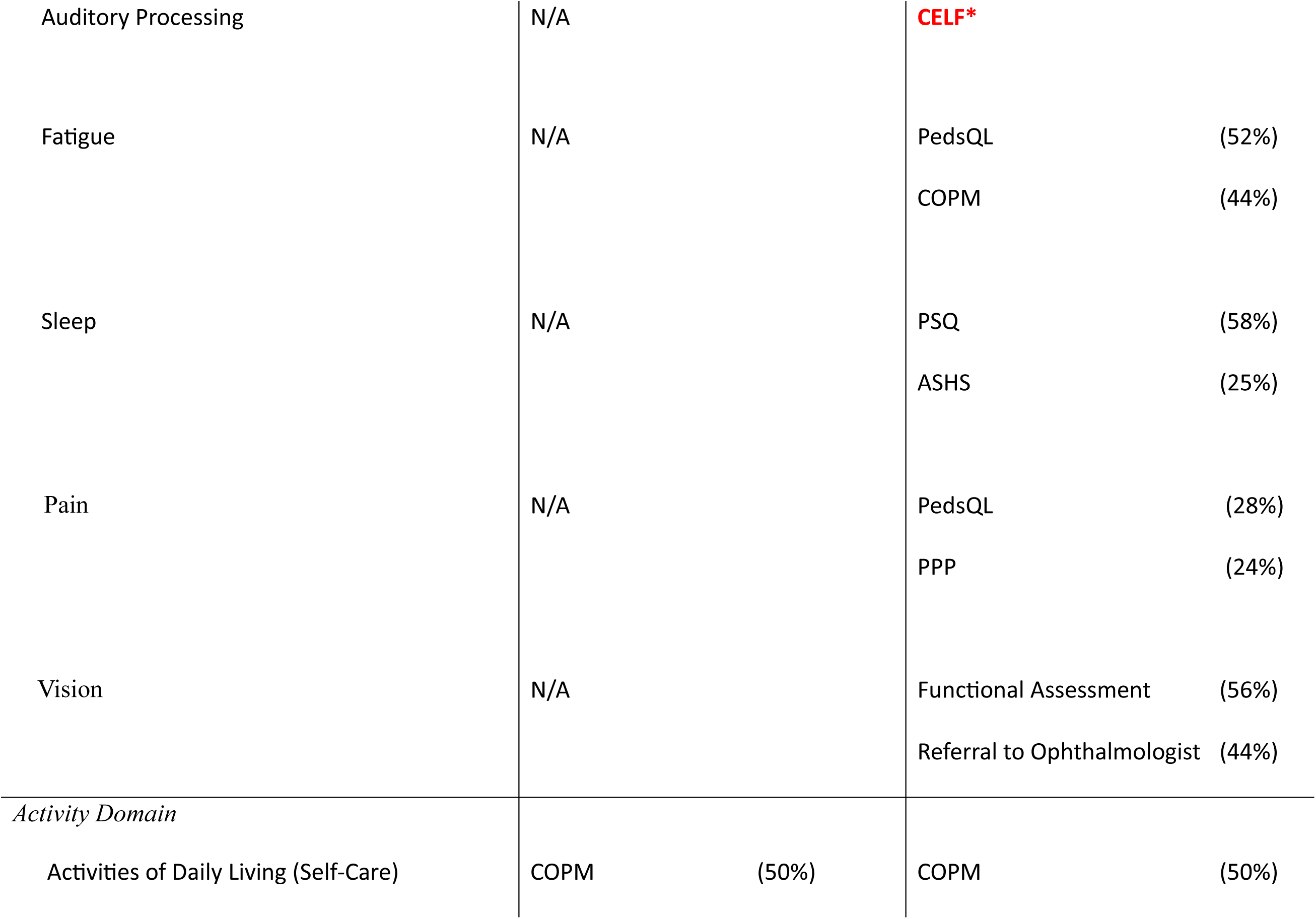
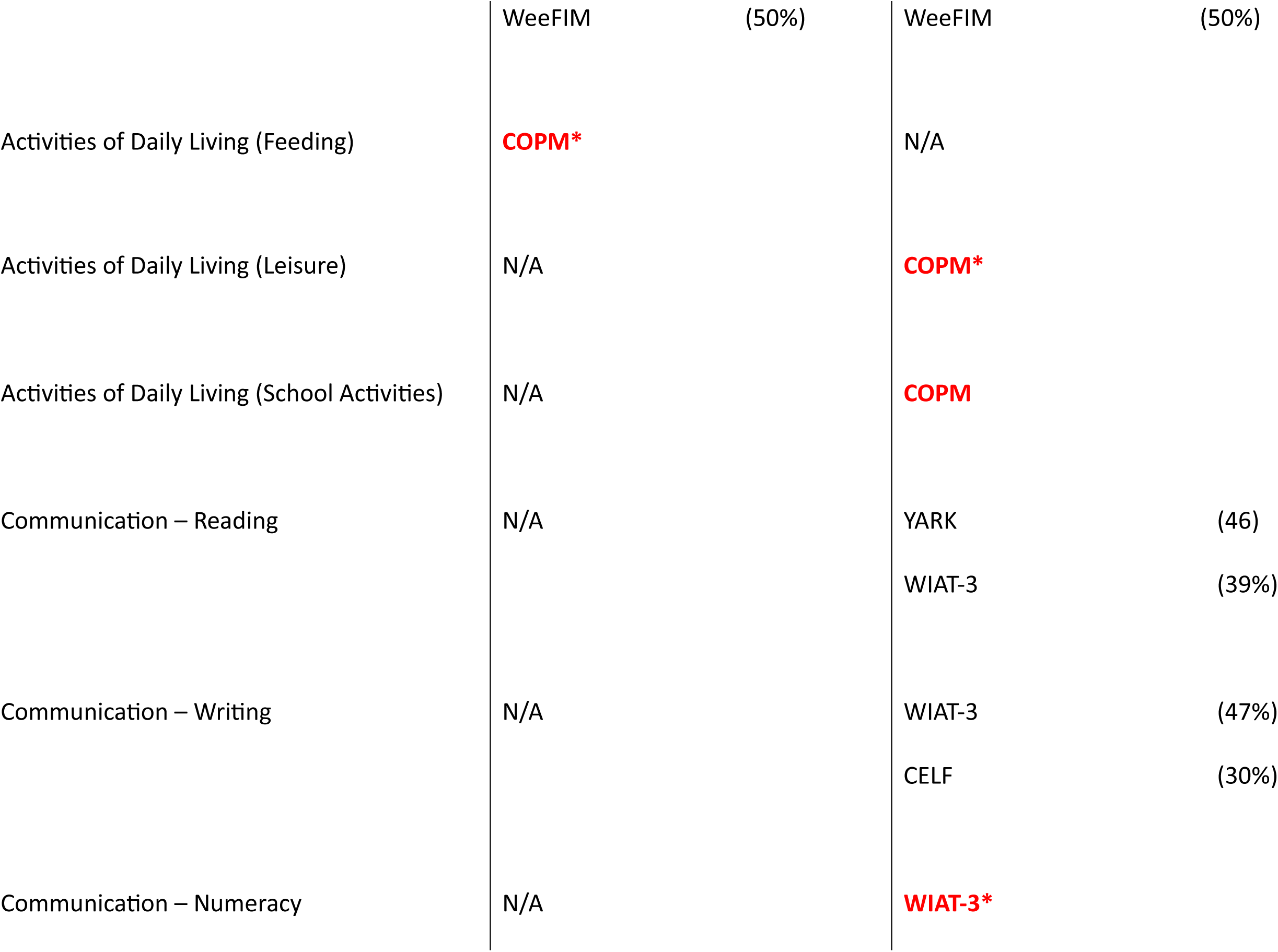
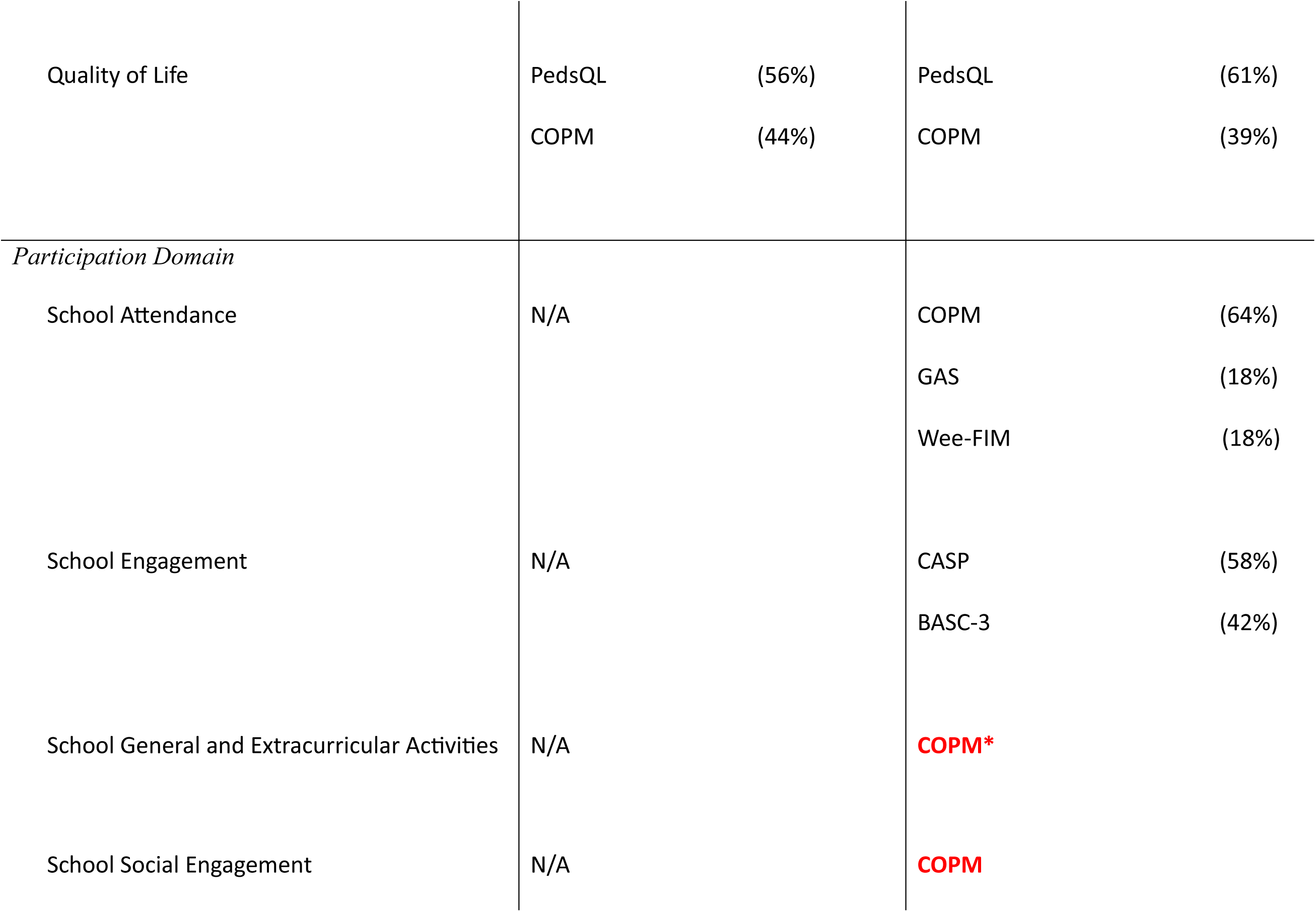
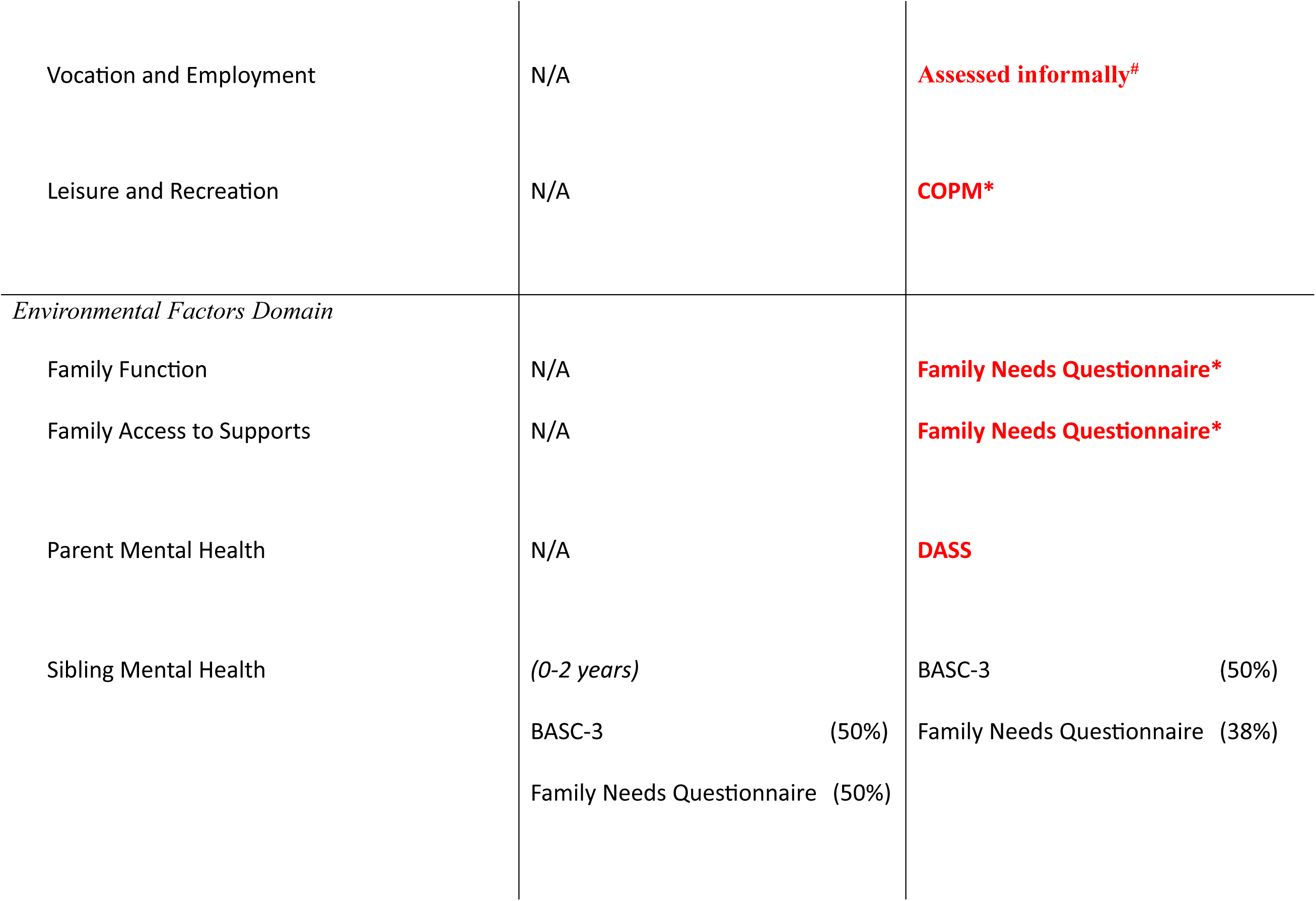
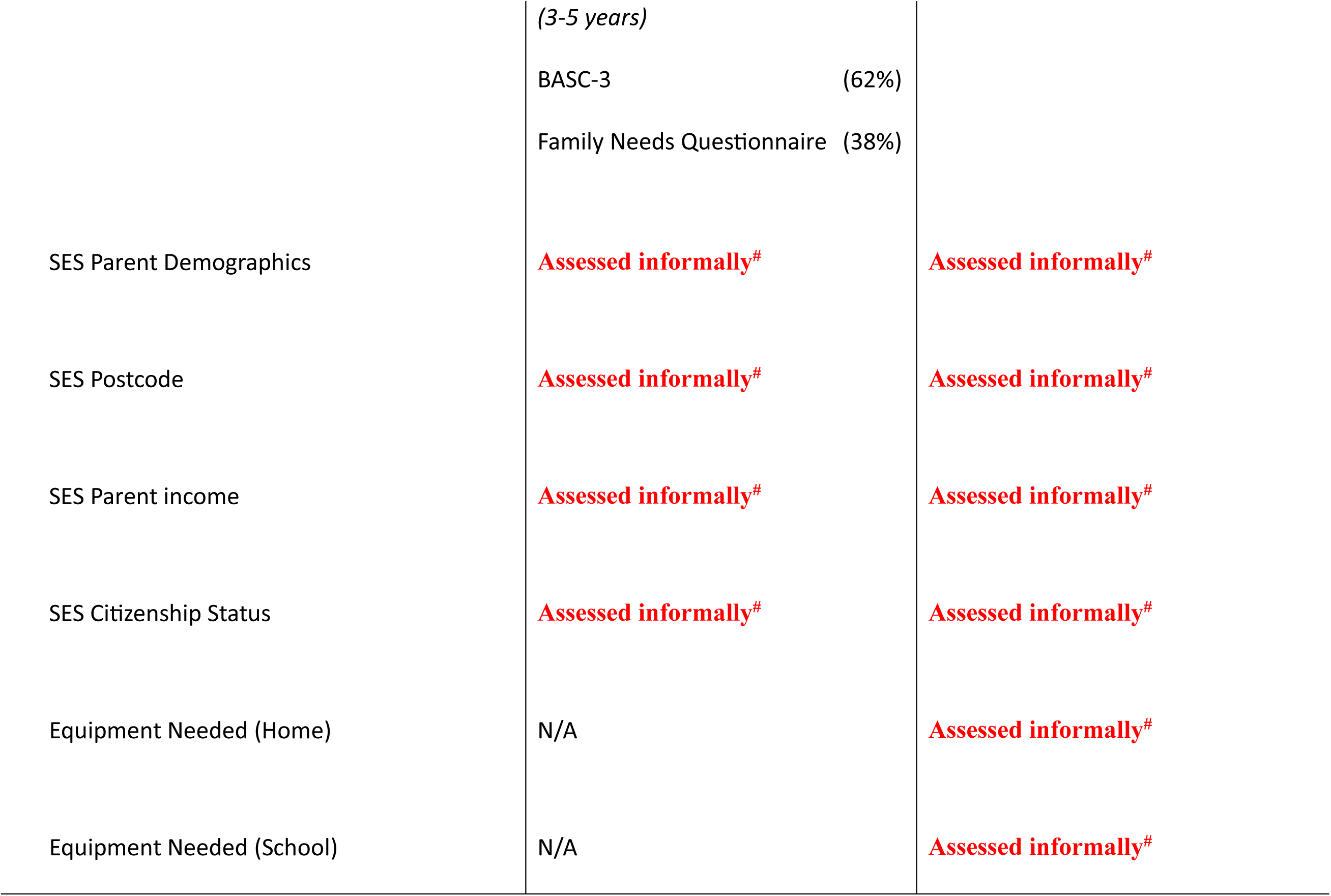
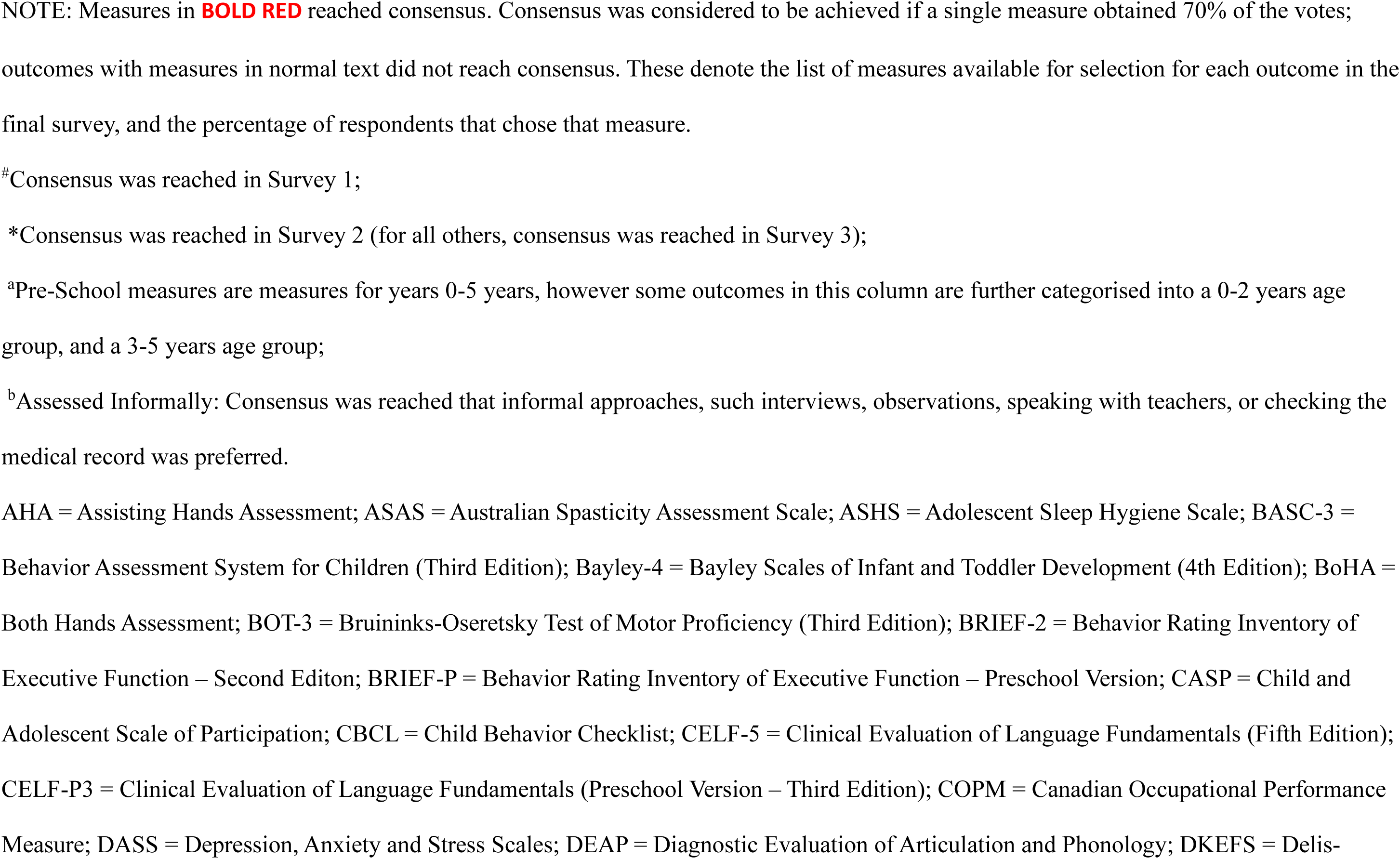
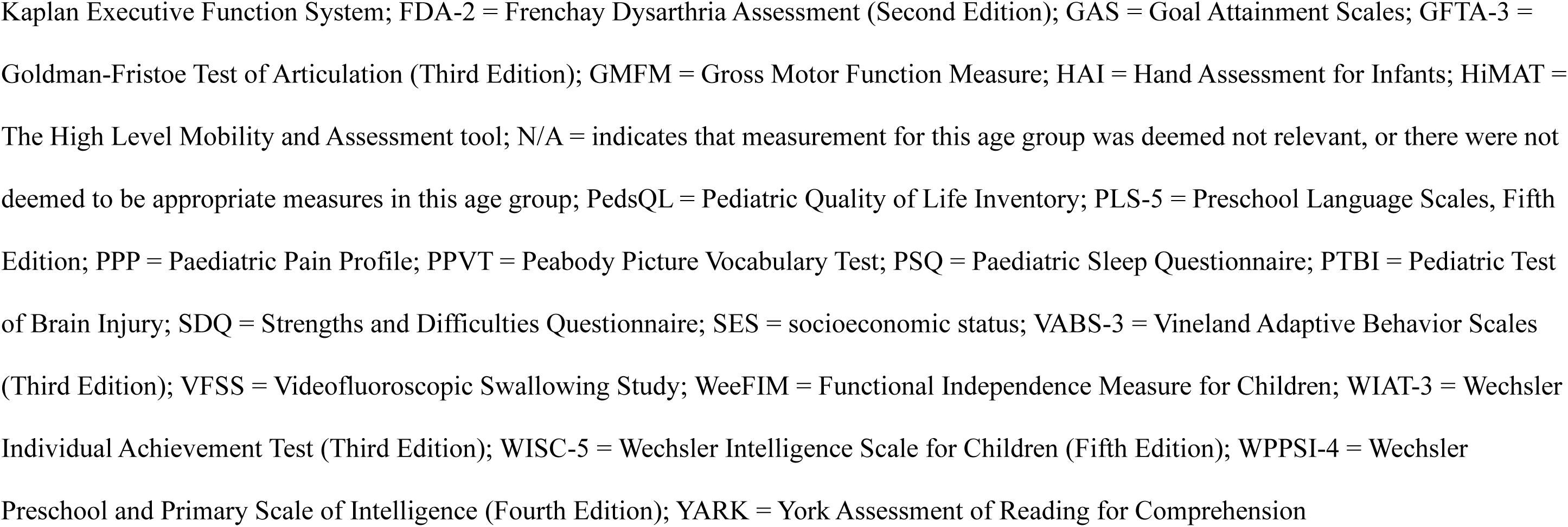
The Outcome Measures for which Consensus was, and was not, reached.

### Surveys 2 and 3

A total of 70 participants provided responses to Survey 2, whereas 57 participants provided responses to Survey 3. Table 4 shows the measures in which consensus was reached regarding the best tools to assess outcomes. The measures that reached consensus are highlighted in BOLD RED. This table shows the findings across all three Surveys.

Consensus was reached for a further 10 measures following Survey 2 (in addition to the 7 in Survey 1). At the conclusion of Survey 3, consensus was reached for a total of 30 outcomes across the different age bands. More specifically, consensus was reached on the measures for 14 (64%) of 22 outcomes in the Pre-School age band, and 20 (50%) of the 40 potential measures in the School Aged age group.

Table 4 also displays the outcomes for which consensus was not reached. For these outcomes, the measures reported in the Table were the top two measures in terms of votes received, and the percentage of votes that each measure obtained. With the cut-off for consensus being 70% agreement, it can be seen that 11 outcomes, including Motor Upper Limb Impairment, Motor Upper Limb Function, Motor Lower Limb Impairment, Receptive Language, Expressive Language, Speech and Speech Sounds, Psychosocial, Emotional and Behaviour Functioning, Cognition, Executive Function, Sleep and School Attendance had measures that received scores that approached the 70% level of agreement. In addition, there were 9 measures, including Receptive Language, Psychosocial, Emotional and Behaviour Functioning, Fatigue, Vision, Activities of Daily Living (Self-Care), Communication (Reading), Quality of Life, School Engagement and Sibling Mental Health, in which there were only two clear measures that received a high proportion of votes, despite not reaching the 70% cut-off for consensus. As a result, consensus was reached for a total of 15 separate measures, across all functional outcomes (see Table 4), with these measures selected to be included in the MDS.

Table 5 displays where consensus was reached for the time points for measurement. The best timing of assessment was not sought for the outcomes that were deemed to be best measured informally (these were all mostly sociodemographic outcomes, which are often found in the medical records and collected at intake). Consensus for the timing of the assessment was reached for every outcome.

**Table 5.**
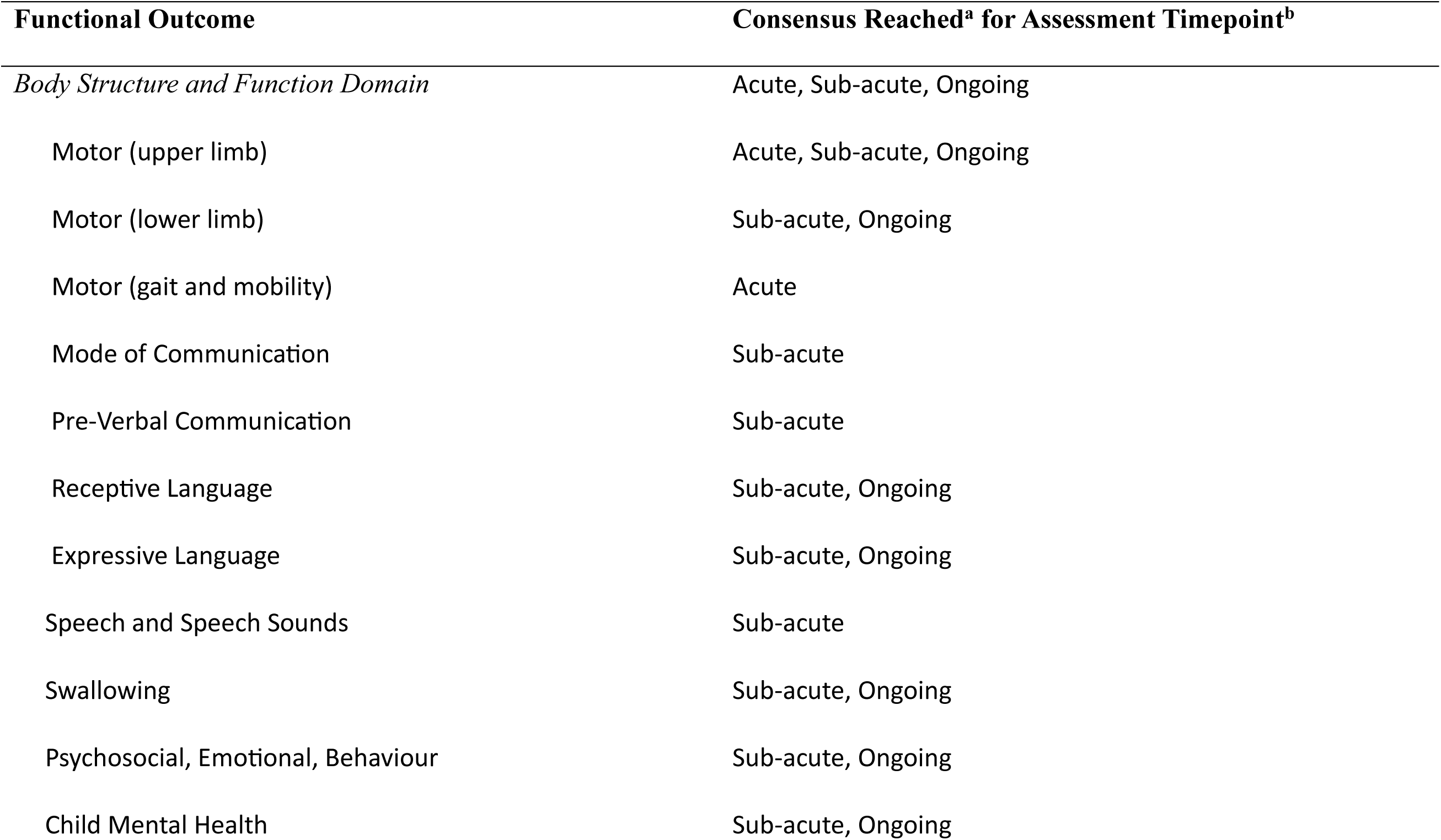
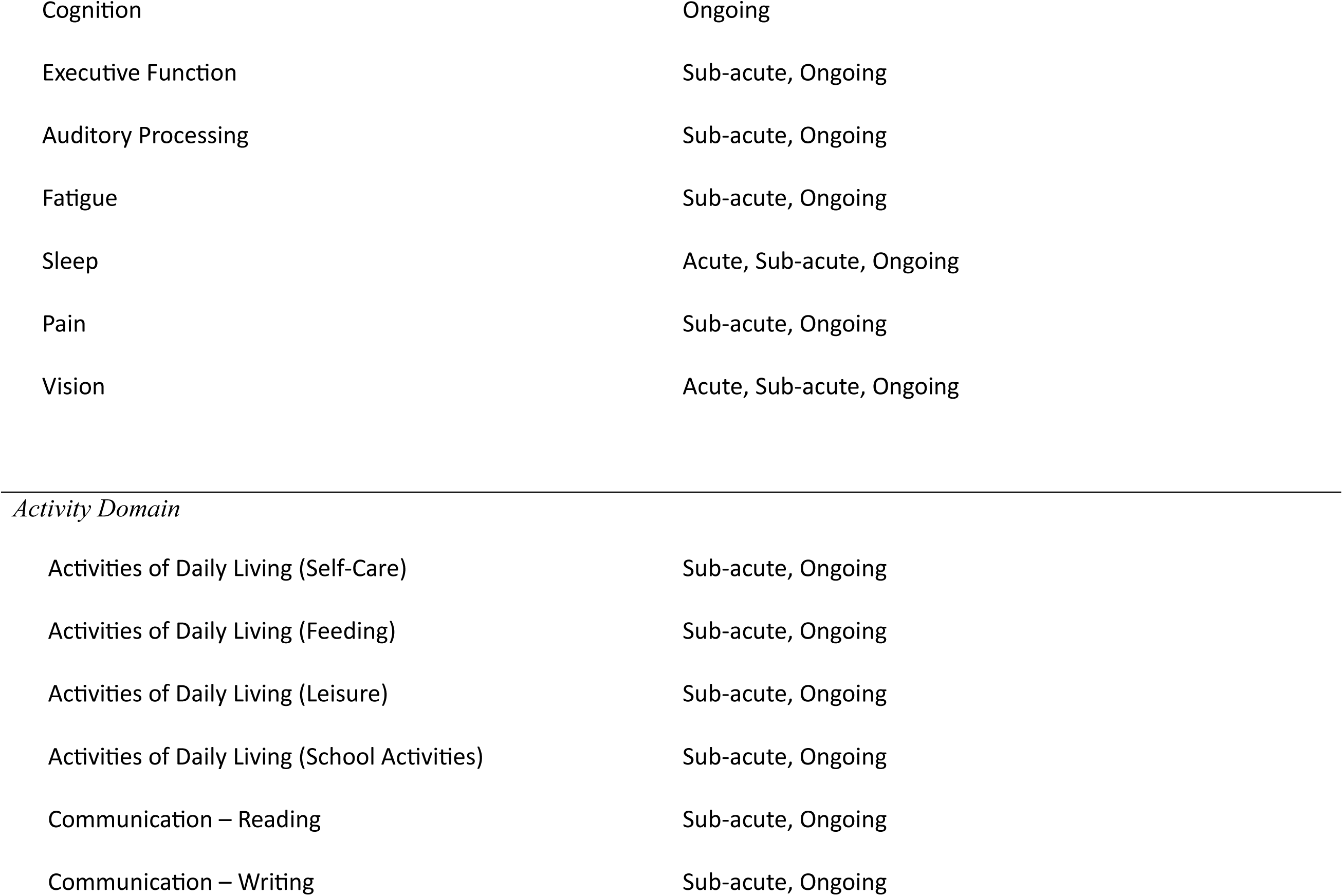
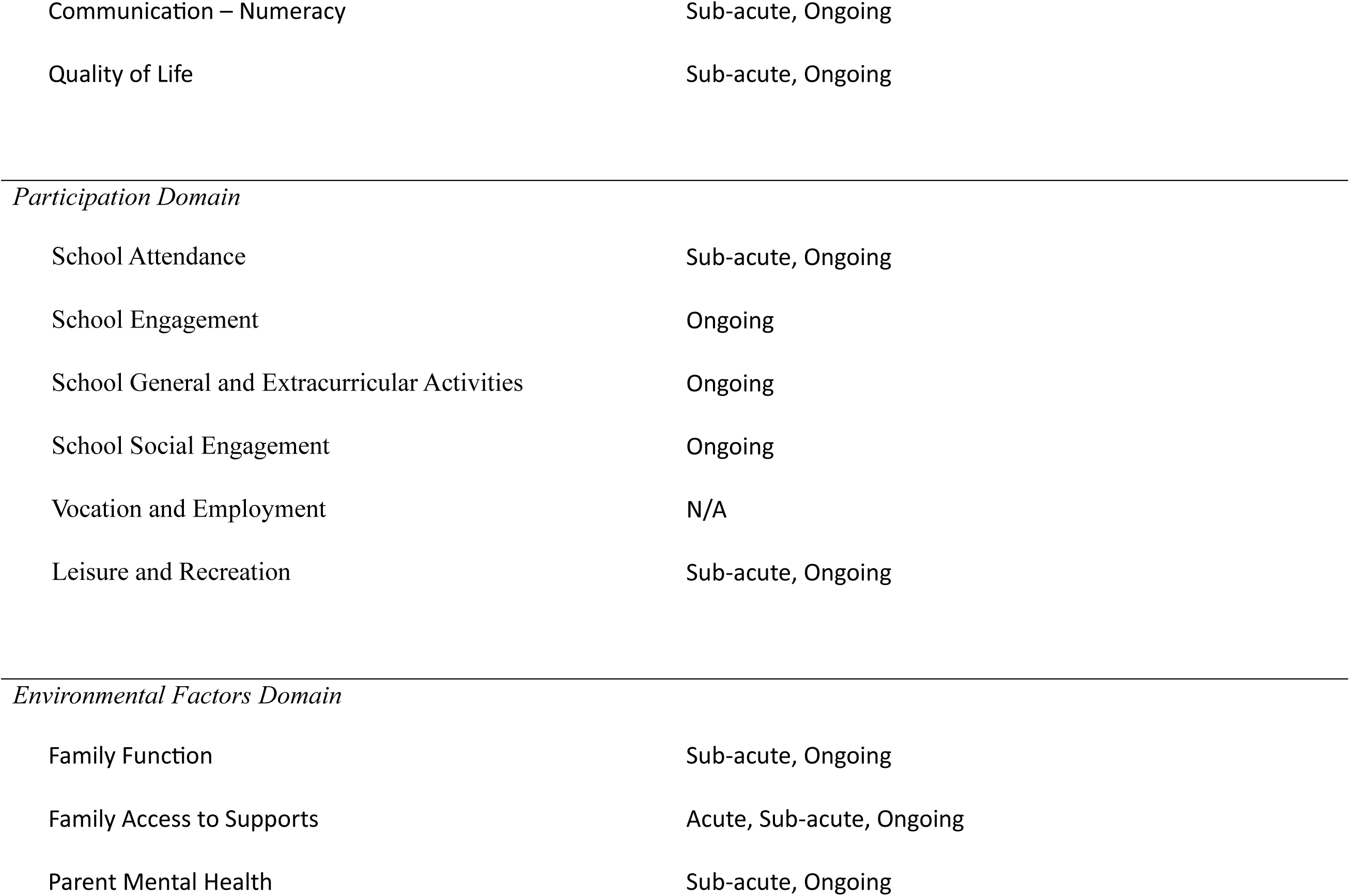
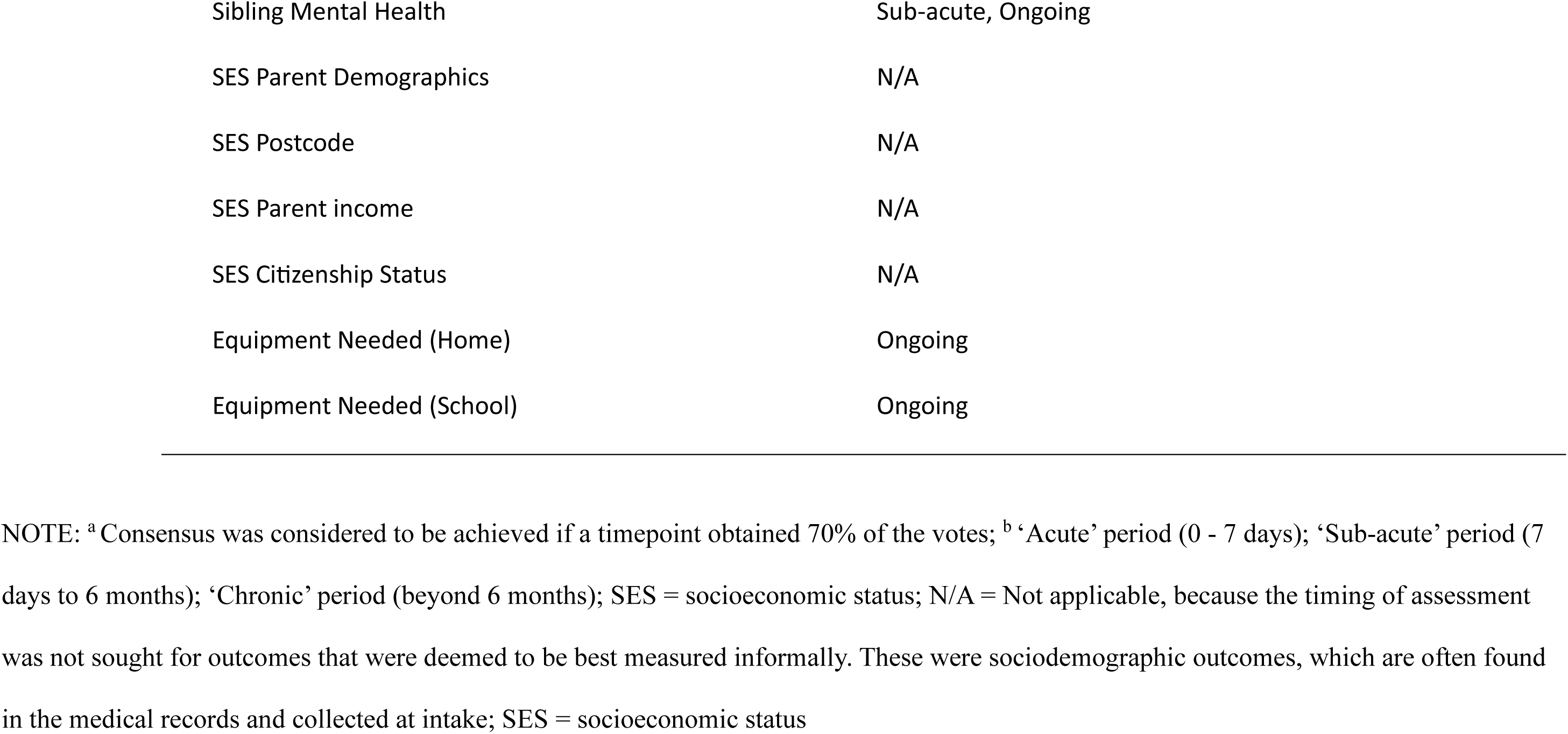
Consensus Regarding the Time Point for Measurement of Each Outcome.

| **Functional Outcome** | **Consensus Reached<sup>a</sup> for Assessment Timepoint<sup>b</sup>** |
| --- | --- |
| *Body Structure and Function Domain* | Acute, Sub-acute, Ongoing |
| Motor (upper limb) | Acute, Sub-acute, Ongoing |
| Motor (lower limb) | Sub-acute, Ongoing |
| Motor (gait and mobility) | Acute |
| Mode of Communication | Sub-acute |
| Pre-Verbal Communication | Sub-acute |
| Receptive Language | Sub-acute, Ongoing |
| Expressive Language | Sub-acute, Ongoing |
| Speech and Speech Sounds | Sub-acute |
| Swallowing | Sub-acute, Ongoing |
| Psychosocial, Emotional, Behaviour | Sub-acute, Ongoing |
| Child Mental Health | Sub-acute, Ongoing |
| Cognition | Ongoing |
| Executive Function | Sub-acute, Ongoing |
| Auditory Processing | Sub-acute, Ongoing |
| Fatigue | Sub-acute, Ongoing |
| Sleep | Acute, Sub-acute, Ongoing |
| Pain | Sub-acute, Ongoing |
| Vision | Acute, Sub-acute, Ongoing |
| *Activity Domain* |  |
| Activities of Daily Living (Self-Care) | Sub-acute, Ongoing |
| Activities of Daily Living (Feeding) | Sub-acute, Ongoing |
| Activities of Daily Living (Leisure) | Sub-acute, Ongoing |
| Activities of Daily Living (School Activities) | Sub-acute, Ongoing |
| Communication – Reading | Sub-acute, Ongoing |
| Communication – Writing | Sub-acute, Ongoing |
| Communication – Numeracy | Sub-acute, Ongoing |
| Quality of Life | Sub-acute, Ongoing |
| *Participation Domain* |  |
| School Attendance | Sub-acute, Ongoing |
| School Engagement | Ongoing |
| School General and Extracurricular Activities | Ongoing |
| School Social Engagement | Ongoing |
| Vocation and Employment | N/A |
| Leisure and Recreation | Sub-acute, Ongoing |
| *Environmental Factors Domain* |  |
| Family Function | Sub-acute, Ongoing |
| Family Access to Supports | Acute, Sub-acute, Ongoing |
| Parent Mental Health | Sub-acute, Ongoing |
| Sibling Mental Health | Sub-acute, Ongoing |
| SES Parent Demographics | N/A |
| SES Postcode | N/A |
| SES Parent income | N/A |
| SES Citizenship Status | N/A |
| Equipment Needed (Home) | Ongoing |
| Equipment Needed (School) | Ongoing |

NOTE: <sup>a</sup> Consensus was considered to be achieved if a timepoint obtained 70% of the votes; <sup>b</sup> ‘Acute’ period (0 - 7 days); ‘Sub-acute’ period (7 days to 6 months); ‘Chronic’ period (beyond 6 months); SES = socioeconomic status; N/A = Not applicable, because the timing of assessment was not sought for outcomes that were deemed to be best measured informally. These were sociodemographic outcomes, which are often found in the medical records and collected at intake; SES = socioeconomic status

## DISCUSSION

Through a Delphi process, this study established a MDS of broad functional outcomes, and appropriate time points to measure them. These outcomes were identified as meaningful to young people with stroke, their families, and clinicians. This study provides an important first step in addressing critical gaps in paediatric stroke rehabilitation research by providing an opportunity to reduce heterogeneity in measurement, and to enhance comparability of data across numerous sites across Australia and New Zealand, with a view to subsequently strengthen the evidence base.

Two contrasting patterns emerged from the Delphi process. There were several functional outcomes where consensus was reached quickly and emphatically. In contrast, agreement on other assessments was less consistent. Body function outcomes, in particular, reached lower levels of consensus, reflecting the availability of multiple tools with differing purposes, as well as a variation in the measurement preferences across different rehabilitation disciplines. Further rounds were required to reach consensus for these functional outcomes, with some not reaching consensus at all after three surveys.

Consensus was reached for 13 of 22 (59%) functional outcomes in the Pre-School age group, and 16 of 36 (42%) measures in the School-Aged age group. Interestingly, consensus was reached quickly (i.e. in Survey 1) for all outcomes where informal assessment was preferred. This is likely due numerous reasons, including the fact that for many of those outcomes, 1) no specific, validated measures exist to measure them (e.g. citizenship status, or vocation); 2) the outcome is a simple figure, or piece of information that does not need measurement (e.g. postcode or parent income) or 3) this outcome is most appropriately collected using informal approaches such as through interviews or observation (e.g. assistive equipment needed at home).

In addition, the COPM was identified as a particularly valuable tool, achieving broad consensus as the most appropriate and feasible measure for several functional outcomes. This is likely due to its clinical utility, flexibility, and alignment with rehabilitation outcomes of importance to families and clinicians. Early consensus was reached in the second round based on the use of the COPM as a measure for several outcomes within both the Activity and Participation domains. This finding is significant as these were areas of functional importance for families, as identified in the LEAG consultation process. Interestingly, the COPM is already routinely collected in the Australasian Rehabilitation Outcomes Centre (AROC) Database (30), which collects basic outcome information following stroke.

There was more variability in responses, and a reduced level of consensus, for the outcomes related to the Body Structure and Function Domain. Specifically, there was a lack of consensus in the Upper and Lower Limb Impairment outcomes, and some speech and language outcomes, particularly for school-aged children. Similarly, there was a lack of consensus in the areas of fatigue, sleep, pain, vision and quality of life. One reason for a reduced level of consensus in the Body Structure and Function outcomes may be the large variety of measures available. The greater level of consensus in the pre-school aged measures is likely due to the fact that there are fewer available measures for use in this age group. In addition, due to the developmental context, younger children have fewer skills and functions that have developed, meaning there are fewer functions that are able to be reliably and validly assessed.

Many of the measures that reached consensus were also listed as important measures in a previous study, which proposed an agreed-upon comprehensive tool kit of measures for evaluation of functional outcomes following childhood stroke (21). Many of the measures included in that study are commonly used by rehabilitation clinicians, and a total of nine measures that reached consensus in this study, were also recommended (21). These measures included the GMFM, AHA, HAI, WeeFIM, BASC-3, WIAT-3, BRIEF-2, WISC-5 and the CELF. This suggests that the measures which reached consensus in the current study have a high level of agreed-upon acceptability, validity and reliability. This previous study by Feldman and colleagues (21) did not, however, engage all end-user health professionals providing rehabilitation care, and the study did not confirm whether the measures included in their recommended list assessed outcomes that were of importance to children affected by stroke and their families. The current findings, therefore, add to our understanding by clarifying the feasibility of these measures (i.e. that clinicians currently use, or will actually use, as part of standard care), and confirms that these measures are assessing outcomes that are of importance to families and young people with stroke.

In general, there are several possible reasons for a lack of consensus across other outcomes. The selection of measures for this process was a complex process, with clinicians not necessarily being aware of the variety of measures available for use, or of the psychometric properties such as the reliability and validity of the measures. Further, there was a broad selection of measures that were suggested for many outcomes. For example, for upper motor outcomes, a total of 34 measures were initially proposed by clinicians to measure this outcome in Survey 1 (see Appendix 3). Some of these measures were not relevant or appropriate for the purposes of this study (e.g. not validated measures), and many of them measured different aspects of functioning and impairment. This means that although there were many valid measures that were proposed, the purposes of the measures varied (e.g. some measured function, some measured impairment, some measured functions that were similar but not the exact constructs or functions required to be measured), increasing the difficulty in reaching consensus. This difference in the *purpose* of measures impacted the reaching of consensus in other ways. For example, in the measurement of the Activities of Daily Living (Self-Care) Outcome, both the COPM and WeeFIM were selected and both received high proportions of clinician votes. The challenge is that although these measures are both valid, they are used for different purposes by the rehabilitation clinicians. Specifically, the COPM is used to determine rehabilitation goals, and measures success in achieving them, whereas the WeeFIM measures functional independence. This means that both are valid and appropriate measures, but they are used for different purposes to measure different things.

Finally, consensus was not reached for some functional outcomes due to the different rehabilitation professions responding, and their preferred use of measures. For example, in the Communication (Reading) outcome, the YARK and the CELF were measures mostly selected by speech pathologists in the sample, and the WIAT-3 was preferred by neuropsychologists. For these reasons, this lack of consensus highlights the complexity and challenges in reaching consensus in measurement across multi- or inter- disciplinary teams. For the functional outcomes that did not reach consensus, subsequent steps to reach consensus may require round table discussions with relevant stakeholders, clinicians, and experts in functional measurement to determine the most feasible measure(s) to use within a clinical context.

The current study achieved consensus for the most appropriate timing of assessment for all outcomes. It was agreed that most outcomes would be assessed within the ‘sub-acute’ and/or ‘ongoing’ periods, which is consistent with the model of rehabilitation measuring change over time during the sub-acute period, and over the long-term in the child’s recovery. Vision, sleep, some motor outcomes, and family access to supports were rated as requiring assessment in the acute period, which is consistent with early assessment of critical functions, and ensuring that the family has access to supports as they move through the initial trauma and adjustment following childhood stroke.

### Strengths and Limitations

This study has several strengths. Use of the Delphi method ensured a rigorous, transparent process for achieving consensus among a diverse group of experts. The iterative approach allowed for the integration of opinions from a range of rehabilitation clinicians, allowing for the refinement of the dataset through successive rounds, culminating in consensus on key functional outcomes, tools, and timepoints for data collection. Additionally, the study’s grounding in the ICF framework provides a structured approach for understanding recovery, and measurement of outcomes over time.

This study benefitted from a broad representation of stakeholders, including clinician end-users from diverse rehabilitation disciplines and health consumers with lived experience. This consumer-focused approach ensured that the findings align with the priorities of young people that have suffered stoke, and their caregivers. Such integration of patient and family priorities is crucial for improving the relevance and utility of research findings in clinical practice. The multidisciplinary engagement ensures that the MDS is applicable across various healthcare settings.

Despite these strengths, the study has certain limitations. The inclusion of clinicians as the primary respondents in the survey may have biased the responses, especially given that this clinician group may not have extensive knowledge in specific ICF domains, and exposure to all outcome measures in their day-to-day practice or the latest data surrounding their psychometric properties. In addition, while the Delphi method is highly effective for consensus-building, consensus could not be reached for some functional domains due to the complexity of outcome measurement in a developmental context, and within a multi- or inter-disciplinary setting. Finally, the study’s focus on Australasian rehabilitation teams may limit the generalisability of findings to other health care settings, although the dataset could serve as a model for similar initiatives worldwide.

### Implications for Clinical Practice

The MDS offers a practical framework for standardising outcome measurement across Australasian paediatric stroke rehabilitation centres. Clinically, the agreed-upon tools provide a roadmap for assessing recovery comprehensively, and in a consistent way. For instance, tools that measure cognitive, motor, and language outcomes are integral to capturing the multi-dimensional impact of paediatric stroke. By identifying timepoints for data collection, the MDS also facilitates longitudinal monitoring of recovery, which is critical given that younger children can ‘grow’ into their deficits. It is also important for tailoring interventions that are aligned to the developmental and recovery trajectory of patients. In addition, regarding the functional outcomes where consensus was not reached for a measurement tool, the refined list of preferred tools provides a platform for future discussion and progression towards consensus. Nevertheless, the list provides clinicians with a narrowed list of appropriate measures to use for those functional outcomes.

Importantly the findings also highlight the need for multidisciplinary or interdisciplinary teams in the rehabilitation context for paediatric stroke, rather than transdisciplinary teams, as a transdisciplinary clinician is unlikely to have the skills to manage the range of outcome measures to assess these outcomes, using the MDS proposed.

### Research Implications and Future Directions

Establishing this MDS addresses a foundational need for consistent data collection, as it allows researchers across Australasia to contribute to a unified database, facilitating larger studies with greater statistical power.

Another important direction for future research involves exploring the implementation of the MDS in clinical settings. Studies are needed to assess how well the dataset integrates into routine care and whether it improves the quality of rehabilitation services. Equally critical is investigating the feasibility of implementing a more standardised approach to data collection, its acceptability to clinicians, and the impact on patient outcomes and family satisfaction.

Finally, a key priority is the validation of the selected tools for use in paediatric stroke populations. While these tools were identified based on expert consensus, empirical studies are needed to evaluate their reliability, validity, and responsiveness in the specific context of childhood stroke. Such validation will strengthen the utility of the dataset, and the credibility in both clinical and research settings.

## Conclusion

The development of the MDS in this study represents an important advancement in paediatric stroke rehabilitation research. By providing a framework for standardised outcome measurement, the dataset will provide a foundation for future multinational collaborations and intervention trials aimed at improving evidence-based care for children with stroke.

While challenges remain, particularly regarding tool validation and implementation, and the design of multi-national intervention trials, the current findings provide a foundation for transformative improvements in the care of children recovering from stroke. Future efforts should focus on validating the MDS, expanding its use internationally, and evaluating its impact on clinical and research outcomes, with the goal being to improve the lives of children and families affected by paediatric stroke.

## Data Availability

Data will be made available on request

## Acknowledgements

The project team would like to acknowledge the following people for contributing their experience and expertise to the project: Emily Alderman, Vicki A. Anderson, Julie Bernhardt, Samudragupta Bora, Roslyn N. Boyd, Andrea Burgess, Loren Conway, Sara Coombes, Alex DiPalma, Lauren Ferguson, Sky Fosbrooke, Briar Gillard, Jennifer Lewis, Megan Lombar, Angela Morgan, Leanne Sakzewski, Kirsty Stewart and Damien Teng.

